# Hospitalized patients lack pathogen-specific comorbidity signatures: general vulnerability and case fatality drive differences in mortality

**DOI:** 10.64898/2026.10.27.26366586

**Authors:** Jacob Bahnsen Schmidt, Frederik Boëtius Hertz, Susanne Häussler

## Abstract

**Objectives:** This study aimed to investigate pathogen-specific prevalence, 30-day mortality, and host factors associated with infection in a large tertiary-care hospital cohort, and to evaluate whether comorbidity patterns and machine-learning models can predict individual outcomes.

**Methods:** We conducted a 39-month retrospective cohort study of 16,282 hospitalized patients with positive bacterial cultures at a tertiary-care university hospital. Cox proportional hazards models identified mortality risk factors. Comorbidity-pathogen associations were examined using logistic regression. Five machine-learning classifiers were trained to predict 30-day mortality.

**Results:** *Enterococcus faecium* contributed deaths comparable to *Staphylococcus aureus* and *Escherichia coli* despite markedly lower prevalence, reflecting the highest case fatality rate across all infection sites (adjusted HR 1.91, 95% CI 1.65–2.22). Multidrug resistance or sex were not independently associated with mortality. Comorbidity profiles were strikingly uniform across pathogens and infection sites; only hematologic malignancies were overrepresented in *E. faecium* bloodstream infections. Machine-learning models achieved limited predictive performance overall, with higher sensitivity only in elderly patients and those with *E. faecium* infections.

**Conclusions:** Nosocomial mortality is shaped by pathogen-specific case fatality rather than prevalence alone. Uniform host vulnerability profiles constrain individual risk prediction and argue for dedicated *E. faecium* surveillance in tertiary-care settings.

## Introduction

Hospital-acquired bacterial infections remain a leading cause of morbidity and mortality in tertiary-care settings, yet the relative contributions of pathogen identity, infection site, and host characteristics to patient outcomes are incompletely understood ^1^. Hospital mortality statistics are typically reported in aggregate, obscuring whether adverse outcomes are primarily driven by the prevalence of certain pathogens, by pathogen-specific case fatality rates, or by underlying host vulnerability. Disentangling these factors is essential for designing targeted prevention strategies and for determining whether individual risk prediction is feasible in heterogeneous inpatient populations.^2–7^

Bacterial pathogens differ in their epidemiology and pathogenic potential, ranging from common causes of infection across diverse populations to opportunistic species primarily affecting immunocompromised or comorbid hosts. In tertiary-care hospitals, both high-prevalence and highly opportunistic pathogens may contribute substantially to mortality, yet whether hospital mortality is predominantly driven by pathogen prevalence or by pathogen-specific case fatality has not been systematically examined in large, unselected inpatient cohorts.

Although numerous studies have examined individual pathogens or specific infection syndromes, integrated analyses combining pathogen identity, infection site, care setting, and host comorbidity in a hospital-wide framework remain scarce. Critically, it is unknown whether patients infected with different pathogens or at different anatomical sites carry distinct comorbidity profiles, a prerequisite for meaningful risk stratification. If comorbidity patterns are largely uniform across infections, this would constrain the predictive value of models relying on pathogen identity, isolation site, and standard clinical variables.

In this study, we analyzed more than 16,000 hospitalized patients over a 39-month period at a tertiary-care university hospital. We systematically examined pathogen prevalence, infection site distribution, and 30-day mortality to determine whether hospital mortality is predominantly driven by pathogen prevalence or by pathogen-specific case fatality. We then compared host comorbidity patterns across pathogens and infection sites to assess whether distinct risk profiles exist, and evaluated whether routinely available clinical and microbiological data allow reliable prediction of individual mortality using machine-learning models.

## Methods

### Data sources

We conducted a retrospective observational study including all patients admitted to Rigshospitalet, Copenhagen, from whom at least one bacterial isolate was recovered between April 1, 2022, and June 16, 2025. As isolation alone does not distinguish infection from colonization, we used antibiotic treatment as a proxy for clinically significant infection in sensitivity analyses.

The primary endpoint was all-cause 30-day mortality, ascertained from patient records. The dataset integrates patient-level information from electronic health records (Sundhedsplatformen, Epic Systems Corporation) and microbiological isolate data, encompassing demographics (age, sex), antibiotic treatment, diagnostic history, and department admission records. Microbiological data included the bacterial species, site of isolation, and multidrug-resistant organism (MDRO) status obtained from the department for Clinical Microbiology database the MADS system (Mikrobiologisk Afdelings Data System). MDRO status was determined per patient and infection type; if any isolate from the same site was an MDRO, the patient was classified as MDRO-positive. Subsequent isolates of the same species from the same anatomical site were excluded from analysis. The study was approved by the local data protection agency (see Ethical approval).

Antibiotic treatment status was assigned if the patient received antibiotics from two days before to seven days after isolate recovery. Patients were classified as ICU patients if the isolate was recovered during an ICU stay. Each ward was grouped into predefined categories (see supplementary file sampling_department_name_categories.xlsx).

Comorbidity diagnoses were extracted from ICD-10 codes recorded prior to or during the index admission; absence of a code was interpreted as absence of the condition. Thirty-day vital status was available for all patients through the Danish Civil Registration System, ensuring complete follow-up. No imputation of missing data was performed.

### Analysis

Statistical analyses were performed using R version 4.2.2. Cox proportional hazards models were fitted to compare hazards associated with different bacterial species, adjusting for age, sex, and comorbidities. p-values for explanatory variables were obtained from partial likelihood ratio tests; p-values for individual levels were derived from Wald tests against the reference category. Associations between comorbidities and pathogen-specific infections were examined using logistic regression in a one-vs-rest setting, comparing each species against all others while adjusting for 30-day outcome. Diagnoses were aggregated into broader categories based on the WHO ICD-10 classification system.

### Patient level 30-day mortality prediction

Using the same covariates as for the Cox models, we trained five machine-learning (ML) classifiers to predict 30-day mortality: logistic regression, K-nearest neighbors (KNN), LightGBM, XGBoost, and a multi-layer perceptron (MLP; hidden layers 128/64, α = 0.01). All models except the MLP used default hyperparameters. To address class imbalance, we tested two oversampling strategies within a 5-fold cross-validation pipeline: the ADASYN algorithm (imbalanced-learn, default parameters) and a conditional generative adversarial network (CGAN). Each generator produced a balanced synthetic training set of 25,000 samples per outcome class. Models were evaluated using sensitivity as the primary metric, alongside accuracy, specificity, F1-score, ROC AUC, and log loss. ML analyses were performed in Python 3 using scikit-learn, LightGBM, XGBoost, and TensorFlow/Keras.

## Results

### Prevalence of bacterial pathogens in hospitalized patients

We analyzed 16,282 hospitalized patients from whom at least one bacterial isolate was recovered over a 39-month period at Rigshospitalet, Copenhagen, Denmark. *Staphylococcus aureus* was the most frequently recovered pathogen, followed by *Escherichia coli*, *S. epidermidis*, *Enterococcus faecalis*, and *E. faecium*. The distribution across anatomical sites varied by species (Fig. 1 and Supplementary Table 1). More positive cultures were obtained from males than females, except in urinary tract infections (Supplementary Fig. 1). The overall higher number of bacterial isolates recovered from male patients reflected cohort composition rather than differential pathogen recovery (Supplementary Table 2). A single bacterial species was isolated in 50.2% of patients, two in 20.9%, and four or more in 13.9%.

**Fig. 1.**
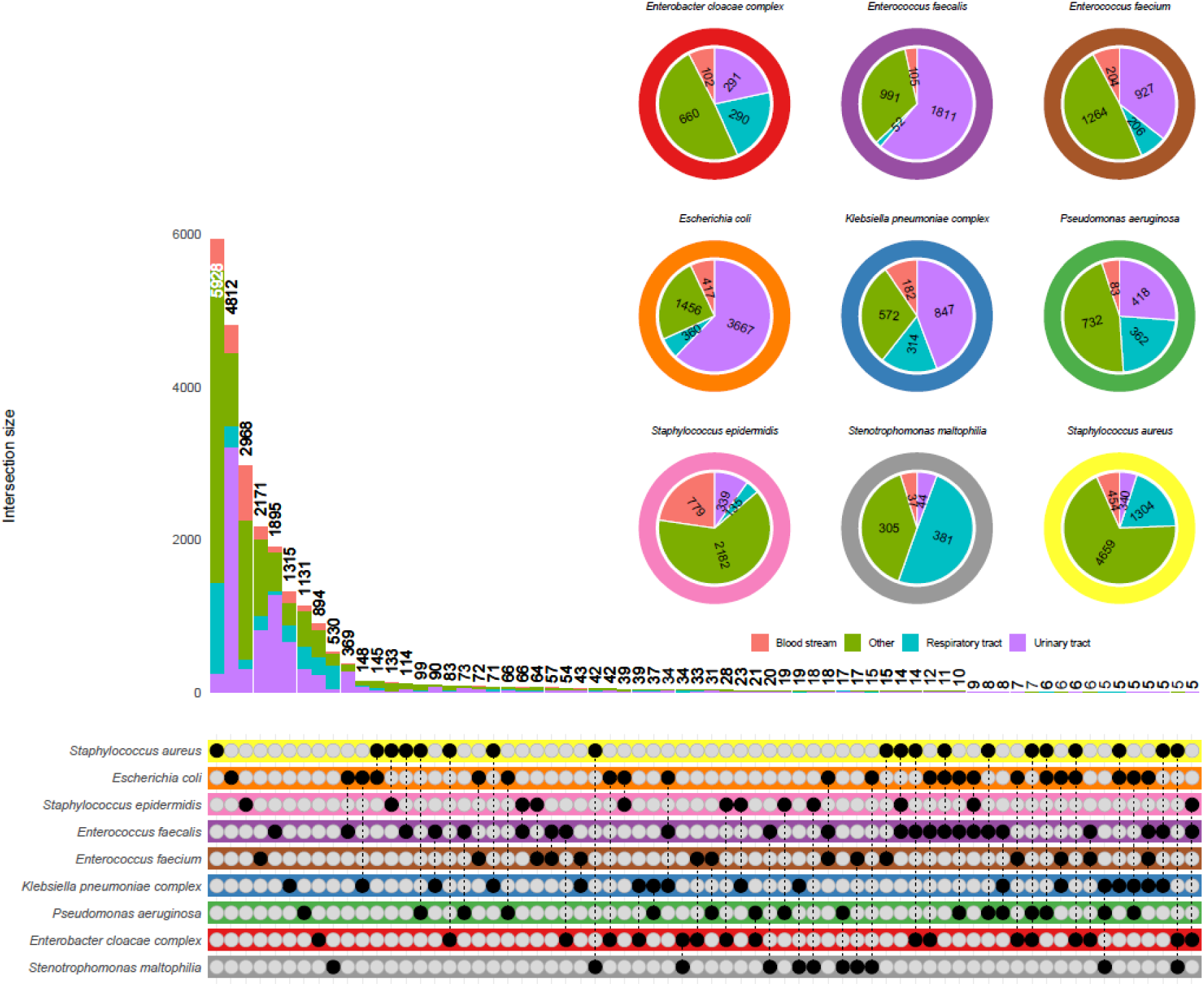
Number of isolates of the nine most frequently identified pathogens at a university hospital, recovered from various infection sites over a 39-month period. The pie charts show the distribution across isolation sites for each species. The UpSet plot (bottom) shows the frequency of multi-species co-isolation patterns.

### 30-day mortality rates associated with the isolation of common pathogens in hospitalized patients

Mortality rates were higher among patients with positive blood cultures or respiratory tract isolates and lower for urinary tract isolates (Fig. 2). Despite its lower prevalence, *E. faecium* contributed a comparable absolute number of deaths as *S. aureus* and *E. coli*, reflecting a substantially higher case fatality rate (Fig. 2 and Supplementary Table 3). *S. maltophilia* exhibited the second-highest mortality rate. Because mortality was attributed to the species isolated prior to death, and multiple isolates were sometimes recovered from the same patient, we also performed a separate analysis of survival probability restricted to patients with a single isolate (n = 5,868). Survival probability declined most steeply for *E. faecium* and *S. maltophilia* (Fig. 3).

**Fig. 2.**
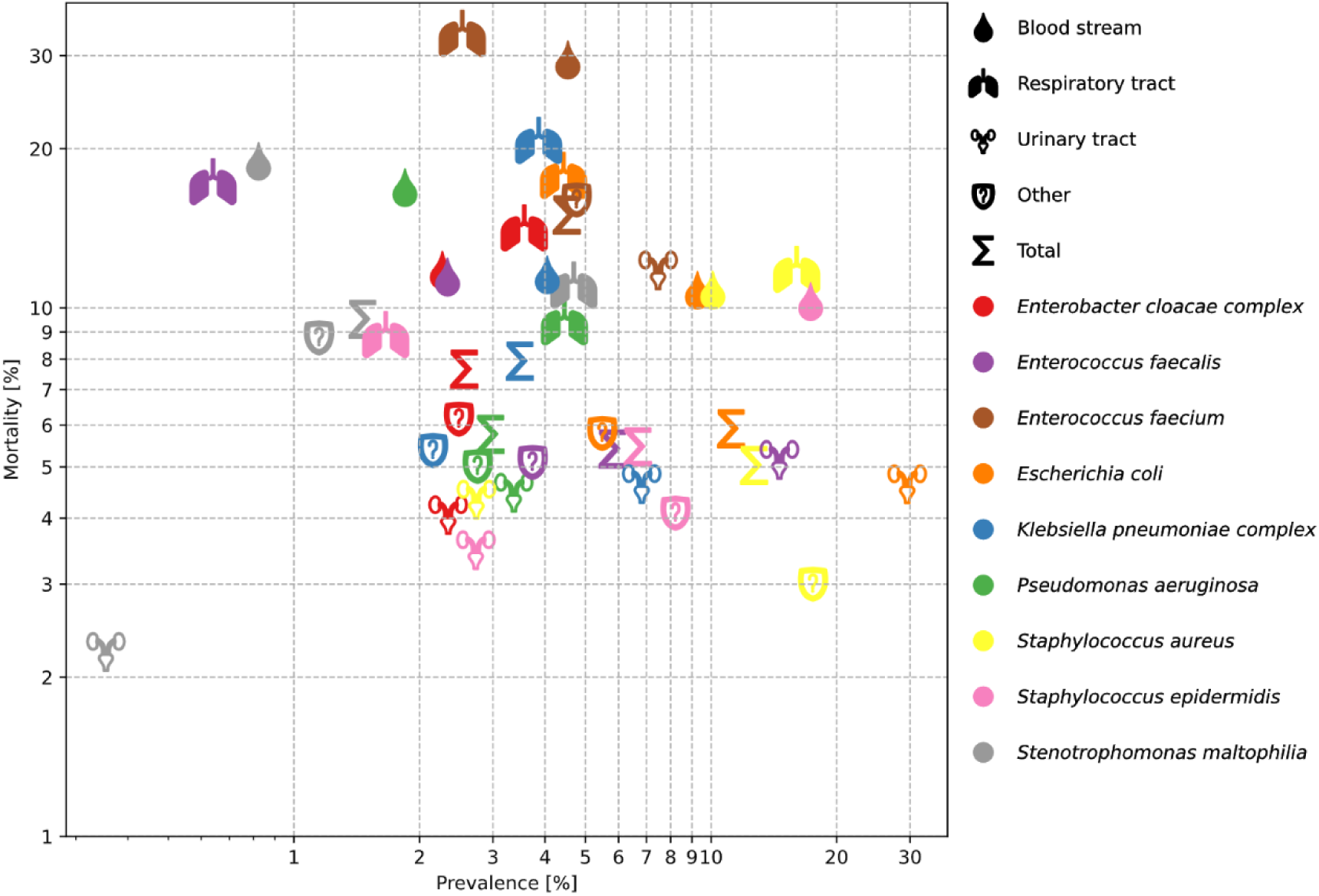
Relationship between 30-day mortality and the prevalence of clinical isolates of various species recovered from patient specimens obtained from different infection sites. Each symbol represents a pathogen-site combination; symbol shape indicates infection site, colour indicates species (see legend).

**Fig. 3.**
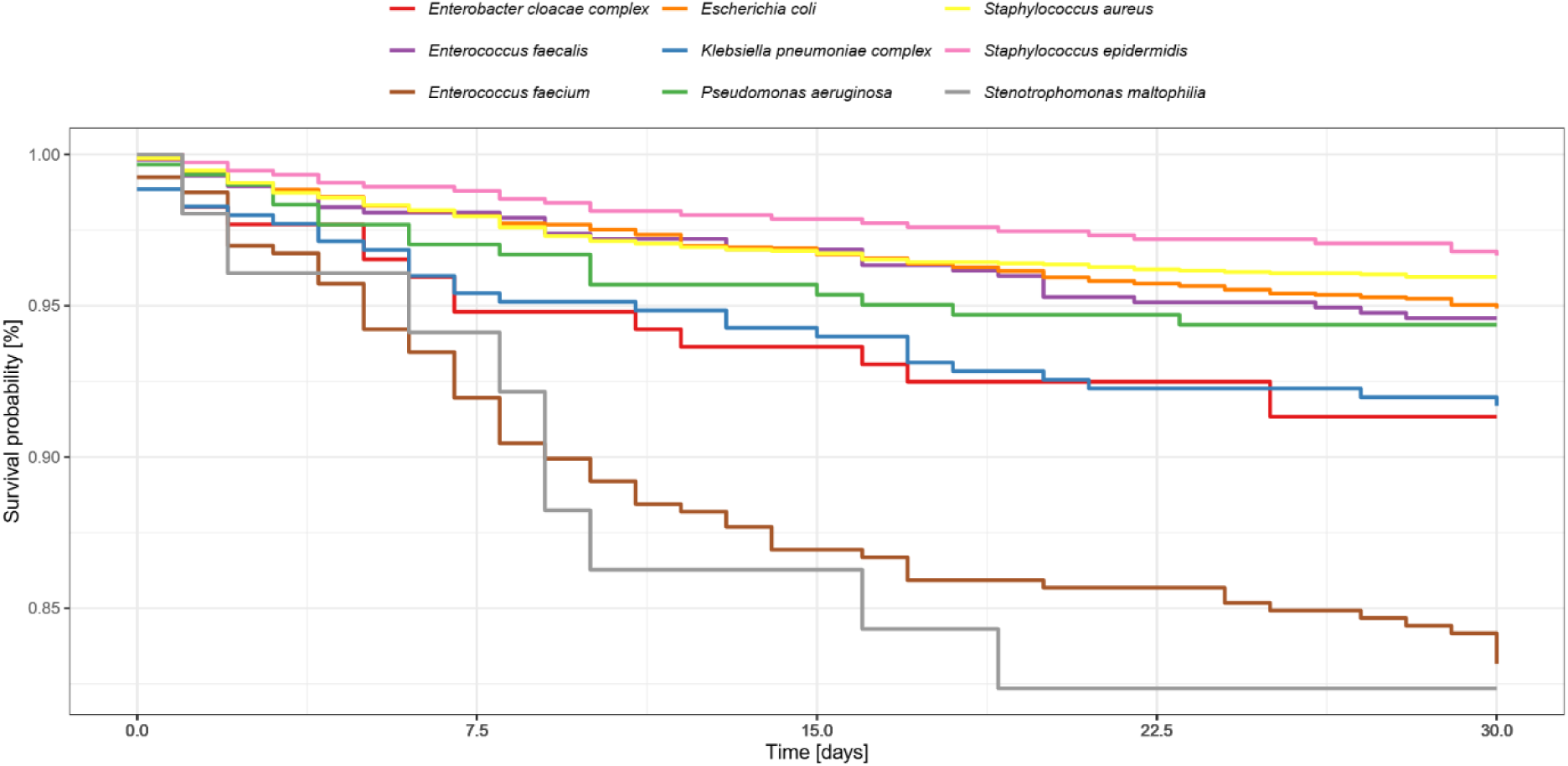
Survival probability (all-cause mortality) as a function of time. Only patients with isolation of a single bacterial species were included in the analysis (n = 5,868).

In an unadjusted Cox proportional hazards model, *E. faecium* and *S. maltophilia* were associated with significantly higher mortality compared with *E. coli* (Table 1). Pathogen recovery from the urinary tract was associated with lower mortality compared with the respiratory tract (HR 0.65; Supplementary Table 4).

**Table 1.** Unadjusted and adjusted hazard ratios (HRs) associated with each bacterial species, compared to *E. coli*. The adjusted HRs were calculated controlling for sex, age, infection site, MDRO status, treatment received, department, ICU admission, preconditions (Charlson comorbidities), and number of pathogens.

| Pathogen | Unadjusted |  | Adjusted |  |
| --- | --- | --- | --- | --- |
|  | HR (95% CI) | p-value | HR (95% CI) | p-value |
| *E. coli* | - | - | - | - |
| *E. cloacae* complex | 1.31 (1.04; 1.65) | **0.024** | 0.94 (0.75; 1.17) | 0.6 |
| *E. faecalis* | 0.91 (0.75; 1.11) | 0.4 | 1.03 (0.85, 1.24) | 0.8 |
| *E. faecium* | 2.67 (2.29; 3.12) | **<0.001** | 1.91 (1.65, 2.22) | **<0.001** |
| *K. pneumoniae* complex | 1.36 (1.11; 1.67) | **0.003** | 1.14 (0.94; 1.38) | 0.2 |
| *P. aeruginosa* | 0.98 (0.77; 1.25) | 0.9 | 0.89 (0.72; 1.07) | 0.3 |
| *S. aureus* | 0.85 (0.73; 1.00) | **0.048** | 0.91 (0.78; 1.07) | 0.2 |
| *S. epidermidis* | 0.92 (0.77; 1.11) | 0.4 | 0.75 (0.62; 0.90) | **0.002** |
| *S. maltophilia* | 1.65 (1.26; 2.15) | **<0.001** | 1.08 (0.83; 1.40) | 0.6 |

Because pathogen detection does not distinguish true infection from colonization, we focused analyses on antibiotic-treated patients, using treatment as a proxy for the clinician’s assessment that the pathogen was causative. Mortality was significantly higher in the treatment group across all infection sites (Table 2; HR 3.21 for treated vs. untreated, Supplementary Table 4). Among treated patients, *E. faecium* infections carried the highest case fatality (Supplementary Fig. 2).

**Table 2.** Number of patients (and 30-day mortality rates) from whom a pathogen was isolated from the airways, the bloodstream or the urinary tract. p-values for the differences in mortality rates between the antibiotic treated and non-treated patient groups were calculated using Pearson’s χ² test.

| Pathogen | Airways |  |  |  | Bloodstream |  |  |  | Urinary tract |  |  |  |
| --- | --- | --- | --- | --- | --- | --- | --- | --- | --- | --- | --- | --- |
|  | Number of isolates (30-day mortality rates in %) |  |  | p-value | Number of isolates (30-day mortality rates in %) |  |  | p-value | Number of isolates (30-day mortality rates in %) |  |  | p-value |
|  | Overall | Treated | Non-treated |  | Overall | Treated | Non-treated |  | Overall | Treated | Non-treated |  |
| *E. coli* | 360<br>(17.8) | 324<br>(18.8) | 36<br>(8.3) | 0.1182 | 417<br>(10.8) | 416<br>(10.6) | 1 (100) | - | 3667<br>(4.7) | 2587<br>(6.1) | 1080<br>(1.3) | <<br>**0.0001** |
| *E. cloacae* complex | 290<br>(14.1) | 259<br>(15.4) | 31<br>(3.2) | 0.0650 | 102<br>(11.8) | 102<br>(11.8) | - | - | 291<br>(4.1) | 231<br>(5.2) | 60 (0.0) | 0.0713 |
| *E. faecalis* | 52<br>(17.3) | 49<br>(16.3) | 3<br>(33.3) | 0.4497 | 105<br>(11.4) | 105<br>(11.4) | - | - | 1811<br>(5.2) | 1372<br>(6.2) | 439<br>(2.3) | **0.0014** |
| *E. faecium* | 206<br>(33.0) | 206<br>(33.0) | - | - | 204<br>(29.4) | 204<br>(29.4) | - | - | 927<br>(12.0) | 874<br>(12.6) | 53 (1.9) | **0.0198** |
| *K. pneumoniae* complex | 314<br>(20.7) | 284<br>(22.2) | 30<br>(6.7) | **0.0461** | 182<br>(11.5) | 180<br>(11.1) | 2<br>(50.0) | - | 847<br>(4.7) | 676<br>(5.5) | 171<br>(1.8) | **0.0405** |
| *P. aeruginosa* | 362<br>(9.4) | 308<br>(10.1) | 54<br>(5.6) | 0.2947 | 83<br>(16.9) | 83<br>(16.9) | - | - | 418<br>(4.5) | 355<br>(5.4) | 63 (0.0) | 0.0602 |
| *S. aureus* | 1304<br>(11.8) | 1012<br>(14.5) | 292<br>(2.4) | **<0.0001** | 454<br>(10.8) | 452<br>(10.6) | 2<br>(50.0) | - | 340<br>(4.4) | 247<br>(5.7) | 93 (1.1) | 0.0660 |
| *S. epidermidis* | 135<br>(8.9) | 121<br>(9.1) | 14<br>(7.1) | 0.8084 | 779<br>(10.3) | 741<br>(10.8) | 38<br>(0.0) | **0.0325** | 339<br>(3.5) | 252<br>(4.0) | 87 (2.3) | 0.4675 |
| *S. maltophilia* | 381<br>(11.0) | 295<br>(13.9) | 86<br>(1.2) | **0.0009** | 37<br>(18.9) | 37<br>(18.9) | - | - | 44<br>(2.3) | 41 (2.4) | 3 (0.0) | - |

### Risk factors associated with mortality in infectious disease patients

Overall mortality did not differ significantly between sexes (Supplementary Table 4). Age was independently associated with mortality (HR 1.04 per year).

Patients from whom multi-drug-resistant organisms (MDROs) were recovered did not experience excess mortality. For *E. faecium*, 30-day mortality was 12.5% among VRE patients (n = 456) compared with 15.4% among non-VRE patients (n = 1,686; difference was not significant, Table 3).

**Table 3.** Number of patients (and 30-day mortality rates) from whom a MDRO or non-MDRO was isolated.

| Bacteria | 30-day mortality |  |  | 90-day mortality |  |  |
| --- | --- | --- | --- | --- | --- | --- |
|  | MDRO | Non-MDRO | p-value | MDRO | Non-MDRO | p-value |
| *E. faecium* | 456<br>(12.5) | 1686<br>(15.4) | 0.094 | 456<br>(20.8) | 1686<br>(23.0) | 0.322 |
| *S. aureus* | 146 (4.1) | 5875<br>(5.0) | 0.606 | 146 (6.2) | 5875<br>(7.3) | 0.566 |
| *E. coli* | 131 (4.6) | 5172<br>(5.9) | 0.516 | 131 (9.9) | 5172<br>(9.9) | 0.961 |
| *K. pneumoniae complex* | 74 (2.7) | 1579<br>(8.2) | 0.089 | 74 (8.1) | 1579<br>(13.0) | 0.209 |

The adjusted HR for MDRO status was 0.92 (Supplementary Table 4), consistent with a previous Danish cohort study. ^8^

Patients who died within 30 days had a higher comorbidity burden (median six vs. four prior diagnoses). Furthermore, ICU admission was strongly associated with mortality (HR 2.20; Supplementary Table 4). Mortality was approximately twice as high for non-ICU *E. faecium* patients compared with non-ICU *E. coli* patients (Supplementary Fig. 3 and Supplementary Table 5; Supplementary Fig. 4). After adjusting for sex, age, infection site, antibiotic treatment, MDRO status, hospital department, Charlson comorbidities, ICU admission, and number of concurrent pathogens (Table 1), *E. faecium* was the only pathogen associated with a significantly higher adjusted hazard ratio compared with *E. coli*. *S. epidermidis* was associated with lower mortality (adjusted HR 0.75, 95% CI: 0.62–0.90; p = 0.002). Individual Charlson comorbidities and departmental hazard ratios are provided in Supplementary Table 4.

### Comorbidity patterns are uniform across pathogens and infection sites

**Error! Reference source not found.**Having established that comorbidity burden is associated with overall mortality, we next asked whether patients who develop infections with different pathogens or at different anatomical sites carry distinct comorbidity profiles, a prerequisite for pathogen- or site-specific risk stratification.

The comorbidity profiles were strikingly homogeneous across all pathogens within each infection site (Fig. 4): regardless of the infecting species, the relative distribution of Charlson comorbidities was virtually identical. Patients who died showed a uniformly higher comorbidity prevalence but the same relative pattern. The only notable exception was the overrepresentation of hematologic malignancies among bloodstream infection patients, particularly for *E. faecium* and *K. pneumoniae*. Among patients who died, those with *E. faecium* infections had a notably higher prevalence of leukemia and lymphoma, and those with *K. pneumoniae* infections were more likely to have congestive heart failure, metastatic solid tumors, and liver disease.

**Fig. 4.**
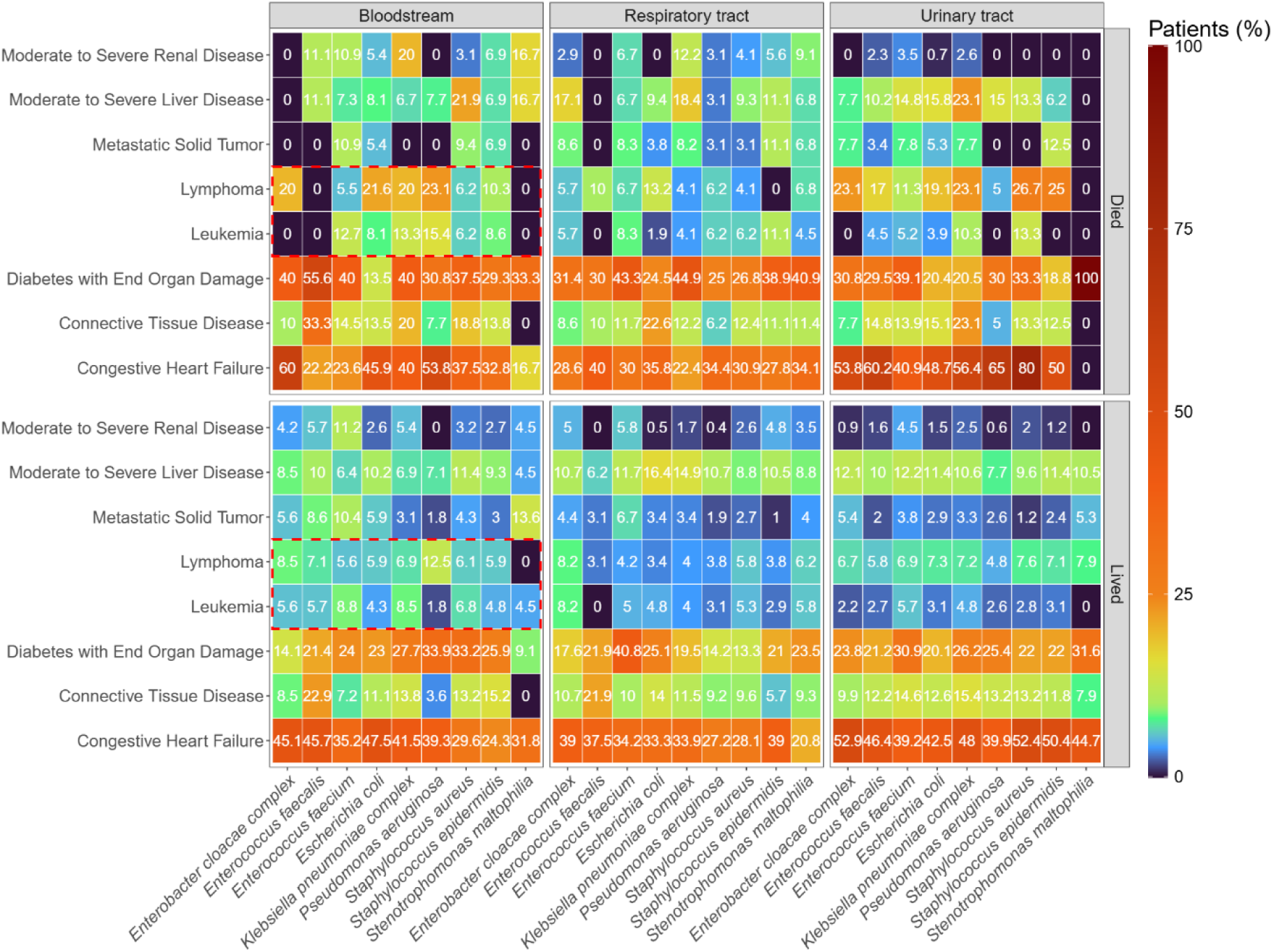
Prevalence of selected Charlson comorbidities across pathogens, stratified by infection site (bloodstream, respiratory tract, urinary tract) and 30-day outcome (survived vs. died). Each cell shows the percentage of patients with a specific comorbidity. Dashed boxes highlight elevated prevalence of hematologic malignancies (lymphoma, leukemia) in bloodstream infection patients.

### Individual mortality prediction is limited by uniform risk architecture

Machine-learning models (logistic regression, KNN, LightGBM, XGBoost, and MLP) demonstrated limited overall performance for 30-day mortality prediction, with sensitivities remaining low even after oversampling to address class imbalance (Supplementary Fig. 5-13). Performance varied markedly across subgroups: sensitivity was highest in elderly patients (≥85 years) and in those with *E. faecium* infections, but remained poor in younger, lower-risk groups (Fig. 5).

**Fig. 5.**
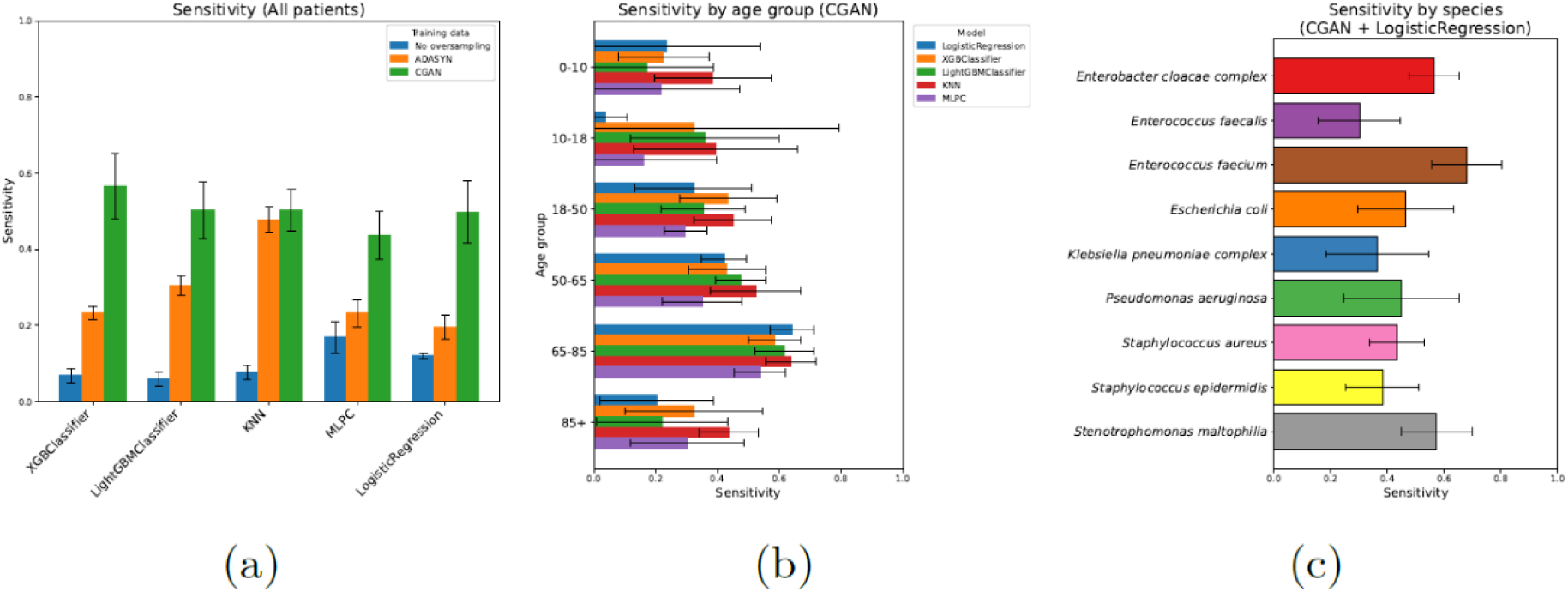
Machine learning model performance for 30-day mortality prediction. (a) Sensitivity of five classification models (XGBClassifier, LightGBMClassifier, K-nearest neighbors, Multilayer perceptron, Logistic Regression) under three oversampling strategies (None, ADASYN, CGAN). (b) Sensitivity by age group (CGAN oversampling). (c) Sensitivity by pathogen (CGAN + Logistic Regression). Error bars represent standard deviations across cross-validation folds.

## Discussion

In this 39-month observational study of more than 16,000 hospitalized patients, recovery of *S. aureus, E. coli,* and *E. faecium* was associated with the highest absolute number of 30-day deaths. Despite its markedly lower prevalence, *E. faecium* contributed a comparable absolute number of deaths, indicating a substantially higher case fatality rate consistent with previous reports highlighting the clinical challenges posed by this pathogen. ^9,10^ This increased mortality was observed irrespective of the site of isolation, and the distribution of *E. faecium* across infection sites did not differ significantly from other major pathogens. Patients with hematologic malignancies were particularly vulnerable to fatal outcomes following *E. faecium* and *S. maltophilia* infections. Conversely, *S. epidermidis* was associated with lower adjusted mortality, likely reflecting that a substantial proportion of positive cultures represent catheter-associated contamination or low-virulence infection in hemodynamically stable patients rather than invasive disease.

Several studies have linked *E. faecium*, particularly vancomycin-resistant enterococci (VRE), to adverse outcomes in nosocomial settings. ^8,11–16^ In our cohort, a considerable proportion of isolates were VRE; however, resistance status was not independently associated with increased mortality. While this contrasts with some reports, it aligns with studies suggesting that host factors and infection severity exert a greater influence on outcome than resistance alone. The absence of a VRE-attributable mortality excess may reflect early identification through active screening rather than a true absence of virulence cost. Nonetheless, the high overall case fatality associated with *E. faecium* regardless of resistance status underscores the importance of surveillance and prevention strategies in vulnerable populations.

Similarly, multidrug resistance in other common pathogens was not independently associated with mortality. Although methicillin-resistant *S. aureus* and multidrug-resistant *E. coli* have been linked to worse outcomes in some studies, the impact of resistance appears highly dependent on patient comorbidities, treatment adequacy, and infection source. ^17–26^ Our findings support the view that resistance alone does not uniformly determine prognosis in heterogeneous hospital populations.

Several studies have reported higher susceptibility to infection and worse outcomes in male patients, particularly in *S. aureus* bacteremia. ^27,28^ In our broad, unselected cohort, however, sex was not independently associated with 30-day mortality after adjustment for age, comorbidities, and infection site. This suggests that previously reported sex differences may be pathogen- or context-specific effects that disappear in heterogeneous hospital populations ^29,30^.

Departmental differences in mortality reflected underlying disease severity, with oncology, hematology, and pediatric departments showing higher hazard ratios, consistent with the case mix of a tertiary referral center. Across all departments, patients who died within 30 days had a broader diagnostic profile, confirming that overall comorbidity burden is a consistent determinant of poor outcome.

To explore whether the clinical and microbiological variables examined here could nonetheless enable individual risk prediction, we evaluated five machine-learning classifiers in an exploratory analysis. Models demonstrated limited overall discriminative performance; although oversampling techniques improved sensitivity, predictive accuracy remained modest. Performance was highest in subpopulations with strong mortality correlates, elderly patients and those with *E. faecium* infections, but declined substantially in lower-risk groups. This subgroup-dependent performance is itself a consequence of the comorbidity uniformity described above: where comorbidity burden is high and distinctive, models found a small signal, whereas in lower-comorbidity groups the uniform host profiles left little discriminatory information.

Our study has several limitations. First, as a single-center study, generalizability to other settings with different patient populations or infection management practices may be limited. Second, despite including over 16,000 patients, residual confounding is possible, particularly regarding unmeasured factors such as timing of therapy, severity scores, and functional status. Third, machine-learning analyses were constrained by variables routinely available in electronic health records and do not capture all determinants of individual mortality. Finally, the observational design precludes causal inference regarding the relative contributions of pathogen, host, and infection site to mortality. Despite these limitations, the large cohort and comprehensive integration of pathogen, site, and host characteristics provide robust insights into infection-related risk patterns in tertiary care.

Taken together, our results support three interconnected conclusions. First, hospital mortality from bacterial infection is driven not only by pathogen prevalence but also by pathogen-specific case fatality: *E. faecium*, despite its relatively low prevalence, contributed a disproportionate mortality burden owing to its high case fatality across all infection sites. Second, the comorbidity profiles of patients were largely uniform across pathogens and infection sites, indicating that susceptibility to nosocomial infection is governed by general host vulnerability rather than by pathogen- or site-specific predisposition. Third, this uniformity explains why individual risk prediction based on routinely available clinical and microbiological data was not achievable in an exploratory machine-learning analysis. These findings have direct implications for clinical practice. Our data suggest that *E. faecium*, despite its lower prevalence, warrants equivalent attention owing to its disproportionate case fatality.

Specifically, we propose that tertiary-care centers consider dedicated *E. faecium* surveillance and management bundles, targeting early identification in high-risk patients, particularly those with hematologic malignancies. More broadly, the absence of pathogen-specific or host signatures implies that risk reduction cannot rely on identifying ‘the right patients’ for targeted interventions, but should instead focus on optimizing care processes, including empirical antibiotic strategies and source control across all patient groups.

## Supporting information

Supplementary figure description

## Data Availability

All relevant data are within the manuscript and its supporting information files. We are unable to share sensitive data instances due to compliance with the General Data Protection Regulation (GDPR) and Danish legislation.

## Author contribution

Conceptualization: Susanne Häussler

Data curation: Jacob Bahnsen Schmidt/ Frederik Boëtius Hertz

Formal analysis: Jacob Bahnsen Schmidt

Investigation: Jacob Bahnsen

Schmidt Methodology: Jacob Bahnsen Schmidt

Ethical approval: Frederik Boëtius Hertz

Resources: Frederik Boëtius Hertz / Jacob Bahnsen Schmidt

Software: Jacob Bahnsen Schmidt

Supervision: Susanne Häussler / Frederik Boëtius Hertz

Validation: Jacob Bahnsen Schmidt

Visualization: Jacob Bahnsen Schmidt

Project administration: Susanne Häussler

Writing- Original Draft Preparation: Jacob Bahnsen Schmidt / Susanne Häussler

Writing – Review & Editing: Susanne Häussler

Materials & Correspondence: Susanne Häussler

## Funding

S.H. was funded by the Novo Nordisk Foundation (NNF 18OC0033946), and received funding from the Deutsche Forschungsgemeinschaft (DFG, German Research Foundation) under Germany’s Excellence Strategy – EXC 2155 “RESIST” – Project ID 390874280, within the SFB/TRR-298-SIIRI – Project-ID 426335750. The funders had no role in designing and conducting the study, analysis, and interpretation of the data, or the writing, review, and approval of the manuscript.

## Ethical approval

There were no ethical concerns for this study. This was an observational, non-intervention study. The extraction of data from registries was approved by the local data protection agency (Journal-nr.: R- 21015888) and registered in Pactius (P-2020-743).

## Code Availability

Access to the code used for the statistical design and ML-process can be made available on reasonable request to the authors by email at

## Competing interest

The authors declare no competing interests.

## Transparency declaration

The lead author affirms that this manuscript is an honest, accurate, and transparent account of the study being reported, that no important aspects of the study have been omitted.

## Notes

### Competing Interest Statement

The authors have declared no competing interest.

### Author Declarations

There were no ethical concerns for this study. This was an observational, non-intervention study. The extraction of data from registries was approved by the local data protection agency (Legal Services, Capital Region of Denmark; Journal Data Center, Center for Health, Capital Region of Denmark; Journal-nr.: R-21015888) and registered in Pactius (P-2020-743).

