## Supplementary tables for "Hospitalized patients lack pathogen-specific comorbidity signatures: general vulnerability and case fatality drive differences in mortality"

*Supplementary Table 1. Distribution of dominant pathogens across different isolation sites*

| Pathogens | Number of isolates from overall 16,282 patients | Number (%) of airway isolates | Number (%) of bloodstream isolates | Number (%) of urinary tract isolates | Percentage of isolates recovered from either of the three isolation sites |
|---|---|---|---|---|---|
| S. aureus | 6021 | 1304 (21.7%) | 454 (7.5%) | 340 (5.6%) | 34.9% |
| *E. coli* | 5303 | 360 (6.8%) | 417 (7.9%) | 3667 (69.1%) | 75.4% |
| *S. epidermidis* | 3171 | 135 (4.3%) | 779 (24.6%) | 339 (19.7%) | 48.6% |
| *E. faecalis* | 2756 | 52 (1.9%) | 105 (3.8%) | 1811 (65.7%) | 71.4% |
| *E. faecium* | 2142 | 206 (9.6%) | 204 (9.5%) | 927 (43.3%) | 62.4% |
| *K. pneumoniae* | 1653 | 314 (19.0%) | 182 (11.0%) | 847 (51.2%) | 81.2% |
| *P. aeruginosa* | 1406 | 362 (25.7%) | 83 (5.9 %) | 418 (29.7%) | 61.3% |
| *E. cloacae complex* | 1216 | 290 (23.8%) | 102 (8.4%) | 291 (23.9%) | 56.1% |
| *S. maltophilia* | 694 | 381 (54.9%) | 37 (5.3%) | 44 (6.3%) | 66.5% |

*Supplementary Table 2a: Differences in sex stratified by bodily site of pathogen isolation. The p-value is from $\chi^2$ test.*

| Bacteria | Bodily site | N male | N female | N died male | N died female | p-value |
|---|---|---|---|---|---|---|
| E. cloacae complex | Airways | 181 (3.8%) | 109 (3.3%) | 25 (13.8%) | 16 (14.7%) | 0.8374 |
| E. cloacae complex | Blood | 64 (2.4%) | 38 (2.1%) | 7 (10.9%) | 5 (13.2%) | 0.7365 |
| E. cloacae complex | Urine | 177 (3.1%) | 114 (1.7%) | 9 (5.1%) | 3 (2.6%) | 0.3043 |
| E. faecalis | Airways | 46 (1.0%) | 6 (0.2%) | 8 (17.4%) | 1 (16.7%) | 0.9648 |
| E. faecalis | Blood | 73 (2.7%) | 32 (1.7%) | 6 (8.2%) | 6 (18.8%) | 0.1185 |
| E. faecalis | Urine | 968 (16.8%) | 843 (12.7%) | 55 (5.7%) | 40 (4.7%) | 0.3724 |
| E. faecium | Airways | 152 (3.2%) | 54 (1.6%) | 53 (34.9%) | 15 (27.8%) | 0.3412 |
| E. faecium | Blood | 129 (4.8%) | 75 (4.1%) | 37 (28.7%) | 23 (30.7%) | 0.7642 |
| E. faecium | Urine | 418 (7.2%) | 509 (7.7%) | 53 (12.7%) | 58 (11.4%) | 0.5489 |
| E. coli | Airways | 250 (5.2%) | 110 (3.3%) | 42 (16.8%) | 22 (20.0%) | 0.4645 |
| E. coli | Blood | 242 (9.1%) | 175 (9.6%) | 24 (9.9%) | 21 (12.0%) | 0.4988 |
| E. coli | Urine | 1205 (20.9%) | 2462 (37.2%) | 55 (4.6%) | 118 (4.8%) | 0.7592 |
| K. pneumoniae complex | Airways | 189 (3.9%) | 125 (3.8%) | 41 (21.7%) | 42 (19.2%) | 0.5935 |

| | | | | | | |
|---|---|---|---|---|---|---|
| K. pneumoniae complex | Blood | 100 (3.8%) | 82 (4.5%) | 13 (13.0%) | 8 (9.8%) | 0.4955 |
| K. pneumoniae complex | Urine | 383 (6.6%) | 464 (7.0%) | 12 (3.1%) | 28 (6.0%) | 0.0476* |
| P. aeruginosa | Airways | 209 (4.3%) | 153 (4.6%9 | 22 (10.5%) | 12 (7.8%) | 0.3873 |
| P. aeruginosa | Blood | 47 (1.8%) | 36 (2.0) | 7 (14.9%) | 7 (19.4%) | 0.5832 |
| P. aeruginosa | Urine | 268 (4.6%) | 150 (2.3%) | 12 (4.5%) | 7 (4.7%) | 0.9291 |
| S. aureus | Airways | 756 (15.7%) | 548 (16.5%) | 94 (12.4%) | 60 (10.9%) | 0.4121 |
| S. aureus | Blood | 276 (10.4%) | 178 (9.7%) | 29 (10.5%) | 20 (11.2%) | 0.8070 |
| S. aureus | Urine | 215 (3.7%) | 125 (1.9%) | 11 (5.1%) | 4 (3.2%) | 0.4067 |
| S. epidermidis | Airways | 77 (1.6%) | 58 (1.7%) | 9 (11.7%) | 3 (5.2%) | 0.1879 |
| S. epidermidis | Blood | 456 (17.1%) | 323 (17.6%) | 53 (11.6%) | 27 (8.4%) | 0.1393 |
| S. epidermidis | Urine | 244 (4.2%) | 95 (1.4%) | 10 (4.1%) | 2 (2.1%) | 0.3724 |
| S. maltophilia | Airways | 215 (4.5%) | 166 (5.0%) | 27 (12.6%) | 15 (9.0%) | 0.2764 |
| S. maltophilia | Blood | 18 (0.7%) | 19 (1.0%) | 3 (16.7%) | 4 (21.1%) | 0.7335 |
| S. maltophilia | Urine | 27 (0.5%) | 17 (0.3%) | 0 (0.0%) | 1 (5.9%) | 0.2024 |

*Supplementary Table 2b: Summary statistics for the number of isolates per patient, stratified by sex and infection site*

| | Sex | min | 1.st quantile | median | mean | 3rd. quantile | max |
|---|---|---|---|---|---|---|---|
| overall | male | 1 | 1 | 2 | 3.9 | 4 | 153 |
| | female | 1 | 1 | 2 | 3.3 | 3 | 286 |
| airways | male | 1 | 1 | 2 | 3.5 | 3 | 61 |
| | female | 1 | 1 | 2 | 4.3 | 4 | 66 |
| bloodstream | male | 1 | 1 | 1 | 2.4 | 3 | 26 |
| | female | 1 | 1 | 1 | 2.4 | 3 | 36 |
| urinary tract | male | 1 | 1 | 1 | 2.6 | 2 | 88 |
| | female | 1 | 1 | 1 | 2.2 | 2 | 243 |
| other | male | 1 | 1 | 2 | 3.2 | 3 | 116 |
| | female | 1 | 1 | 1 | 2.6 | 3 | 101 |

*Supplementary Table 3: Prevalence and mortality rate of selected pathogens*

| | | Overall | BSI | RTI | UTI |
|---|---|---|---|---|---|
| Bacteria | Prevalence N (%) | 30-day mortality N (%) | 30-day mortality N (%) | 30-day mortality N (%) | 30-day mortality N (%) |
| *S. aureus* | 6021 (12.7) | 300 (5.0) | 49 (10.8) | 154 (11.8) | 15 (4.4) |
| *E. coli* | 5303 (11.2) | 311 (5.9) | 45 (10.8) | 64 (17.8) | 173 (4.7) |
| *S. epidermidis* | 3171 (6.7) | 173 (5.5) | 80 (10.3) | 12 (8.9) | 12 (3.5) |
| *E. faecalis* | 2756 (5.8) | 146 (5.3) | 12 (11.4) | 9 (17.3) | 95 (5.2) |
| *E. faecium* | 2142 (4.5) | 316 (14.8) | 60 (29.4) | 68 (33.0) | 111 (12.0) |
| *K. pneumoniae complex* | 1653 (3.5) | 131 (7.9) | 21 (11.5) | 65 (20.7) | 40 (4.7) |
| *P. aeruginosa* | 1406 (3.0) | 81 (5.8) | 14 (16.9) | 34 (9.4) | 19 (4.5) |
| *E. cloacae* | 1216 (2.6) | 91 (7.5) | 12 (11.8) | 41 (14.1) | 12 (4.1) |
| *S. maltophilia* | 694 (1.5) | 66 (9.5) | 7 (18.9%) | 42 (11.0) | 1 (2.3) |

*Supplementary Table 4: Cox proportional hazards model including all single-term confounding factors. For more information on how the departments were grouped, see Supplementary File* sampling_department_names_categories.xlsx.

| Parameter | HR (95% CI) | p-value |
|---|---|---|
| **Infection type** | - | **<0.001** |
| Airways | - | - |
| Bloodstream | 1.10 (0.94; 1.29) | 0.2 |
| Urinary tract | 0.64 (0.56; 0.73) | **<0.001** |
| Other | 0.49 (0.42; 0.58) | **<0.001** |
| **Sex** | - | 0.4 |
| Female | - | - |
| Male | 0.96 (0.87; 1.06) | 0.9 |
| **Received treatment** | - | **<0.001** |
| No | - | - |
| Yes | 3.21 (2.55; 4.04) | **<0.001** |
| **Admitted to ICU** | - | **<0.001** |
| No | - | - |
| Yes | 2.20 (1.57; 3.09) | **<0.001** |

| | | |
|---|---|---|
| **MDRO positive** | - | 0.14 |
| No | - | - |
| Yes | 0.92 (0.74; 1.13) | 0.4 |
| **Department** | - | **<0.001** |
| Anaesthesiology | - | - |
| Cardiology | 0.57 (0.42; 0.76) | **<0.001** |
| Cardiothoracic surgery | 0.98 (0.66; 1.45) | >0.9 |
| Child and adolescent psychiatry | 0.00 (0.00; Inf) | >0.9 |
| Clinical genetics | 0.00 (0.00; Inf) | >0.9 |
| Clinical microbiology | 0.61 (0.22; 1.74) | 0.4 |
| Dermatovenerology | 0.00 (0.00; Inf) | >0.9 |
| Emergency medicine | 0.92 (0.58; 1.45) | 0.7 |
| Endocrinology | 0.47 (0.14; 1.54) | 0.2 |
| Gastroenterology and hepatology | 0.46 (0.28; 0.75) | **0.002** |
| Hematology | 0.58 (0.39; 0.87) | 0.008 |
| Immunology | 0.00 (0.00; Inf) | >0.9 |
| Infectious diseases | 0.41 (0.25; 0.65) | **<0.001** |
| Nephrology | 0.30 (0.20; 0.47) | **<0.001** |
| Neurology | 1.05 (0.72; 1.51) | 0.8 |
| Neurosurgery | 0.56 (0.35; 0.90) | **0.016** |
| Obstetrics and gynecology | 0.24 (0.13; 0.46) | **<0.001** |
| Oncology | 0.96 (0.64; 1.43) | 0.8 |
| Ophthalmology | 0.18 (0.06; 0.52) | **<0.001** |
| Orthopedic surgery | 0.44 (0.29; 0.69) | **<0.001** |
| Otorhinolaryngology | 0.31 (0.16; 0.60) | **<0.001** |
| Pediatrics | 4.73 (3.01; 7.45) | **<0.001** |
| Plastic surgery | 0.37 (0.22; 0.60) | **<0.001** |
| Psychiatry | 0.00 (0.00; Inf) | >0.9 |
| Radiology | 0.00 (0.00; Inf) | >0.9 |
| Respiratory medicine | 0.42 (0.24; 0.73) | **0.002** |
| Rheumatology | 0.22 (0.10; 0.51) | **<0.001** |
| Surgery | 0.22 (0.14; 0.34) | **<0.001** |
| Urology | 0.25 (0.17; 0.39) | **<0.001** |

| | | |
|---|---|---|
| Vascular surgery | 0.29 (0.18; 0.47) | **<0.001** |
| **Age** | 1.04 (1.03; 1.04) | **<0.001** |
| **Myocardial Infarction** | 0.84 (0.68, 1.04) | 0.11 |
| **Congestive Heart Failure** | 1.24 (1.08, 1.43) | **0.003** |
| **Peripheral Vascular Disease** | 0.89 (0.76, 1.04) | 0.13 |
| **Cerebrovascular Disease** | 0.99 (0.88, 1.11) | 0.9 |
| **Dementia** | 0.71 (0.58, 0.86) | **<0.001** |
| **Chronic Pulmonary Disease** | 1.01 (0.89, 1.14) | >0.9 |
| **Connective Tissue Disease** | 1.23 (1.02, 1.48) | **0.027** |
| **Ulcer Disease** | 0.88 (0.71, 1.09) | 0.2 |
| **Mild Liver Disease** | 0.97 (0.77, 1.24) | 0.8 |
| **Diabetes** | 0.85 (0.73, 0.99) | **0.041** |
| **Hemiplegia** | 0.20 (0.13, 0.30) | **<0.001** |
| **Moderate to Severe Renal Disease** | 1.27 (1.13, 1.42) | **<0.001** |
| **Diabetes with End Organ Damage** | 1.26 (1.06, 1.48) | **0.007** |
| **Any Tumor** | 0.93 (0.83, 1.04) | 0.2 |
| **Leukemia** | 1.46 (1.17, 1.83) | **<0.001** |
| **Lymphoma** | 1.47 (1.23, 1.76) | **<0.001** |
| **Moderate to Severe Liver Disease** | 1.70 (1.32, 2.19) | **<0.001** |
| **Metastatic Solid Tumor** | 1.24 (1.05, 1.48) | **0.013** |
| **AIDS** | 0.92 (0.54, 1.58) | 0.8 |
| **N Pathogens** | 1.02 (1.00, 1.05) | **0.033** |

*Supplementary Table 5: Number of ICU and Non-ICU patients from whom a pathogen was recovered and mortality in (%).*

| | 30-day mortality | | 90-day mortality | |
|---|---|---|---|---|
| | ICU | Non-ICU | ICU | Non-ICU |
| *S. aureus* | 484 (23.1) | 5537 (3.4) | 484 (26.9) | 5537 (5.5) |
| *E. coli* | 309 (23.9) | 4994 (4.7) | 309 (29.1) | 4994 (8.7) |
| *S. epidermidis* | 280 (15.0) | 2891 (4.5) | 280 (19.3) | 2891 (8.1) |
| *E. faecalis* | 189 (18.0) | 2567 (4.4) | 189 (25.4) | 2567 (8.8) |
| *E. faecium* | 386 (34.7) | 1756 (10.4) | 384 (43.5) | 1748 (17.9) |
| *K. pneumoniae* | 172 (23.3) | 1481 (6.1) | 172 (27.3) | 1481 (11.1) |
| *P. aeruginosa* | 159 (19.5) | 1247 (4.0) | 159 (25.8) | 1247 (8.3) |
| *E. cloacae* | 180 (17.2) | 1036 (5.8) | 180 (27.8) | 1036 (10.1) |
| *S. maltophilia* | 136 (28.7) | 558 (4.8) | 136 (39.7) | 558 (11.3) |
