## Supplementary figure description for "Hospitalized patients lack pathogen-specific comorbidity signatures: general vulnerability and case fatality drive differences in mortality"

**Supplementary Fig. 1** Number of patients with positive isolates, stratified by recovery site, pathogen species, and sex. The number above each bar indicates the 30-day mortality rate for the corresponding group.

**Supplementary Fig. 2** Relationship between 30-day mortality and the prevalence of clinical isolates, stratified by antibiotic treatment status (treated vs. non-treated).

**Supplementary Fig. 3** Number of patients testing positive for either *E. faecium* (top row) or *E. coli* (bottom row), stratified by department. Each facet differentiates between ICU and non-ICU patients. Colours indicate 30-day survival status following pathogen isolation (blue = survived; red = died).

**Supplementary Fig. 4** Relationship between 30-day mortality and the prevalence of clinical isolates, stratified by ICU vs. non-ICU admission at the time of pathogen isolation.
