## Supplementary figures and images for "Hospitalized patients lack pathogen-specific comorbidity signatures: general vulnerability and case fatality drive differences in mortality"

### Supplementary figure 1

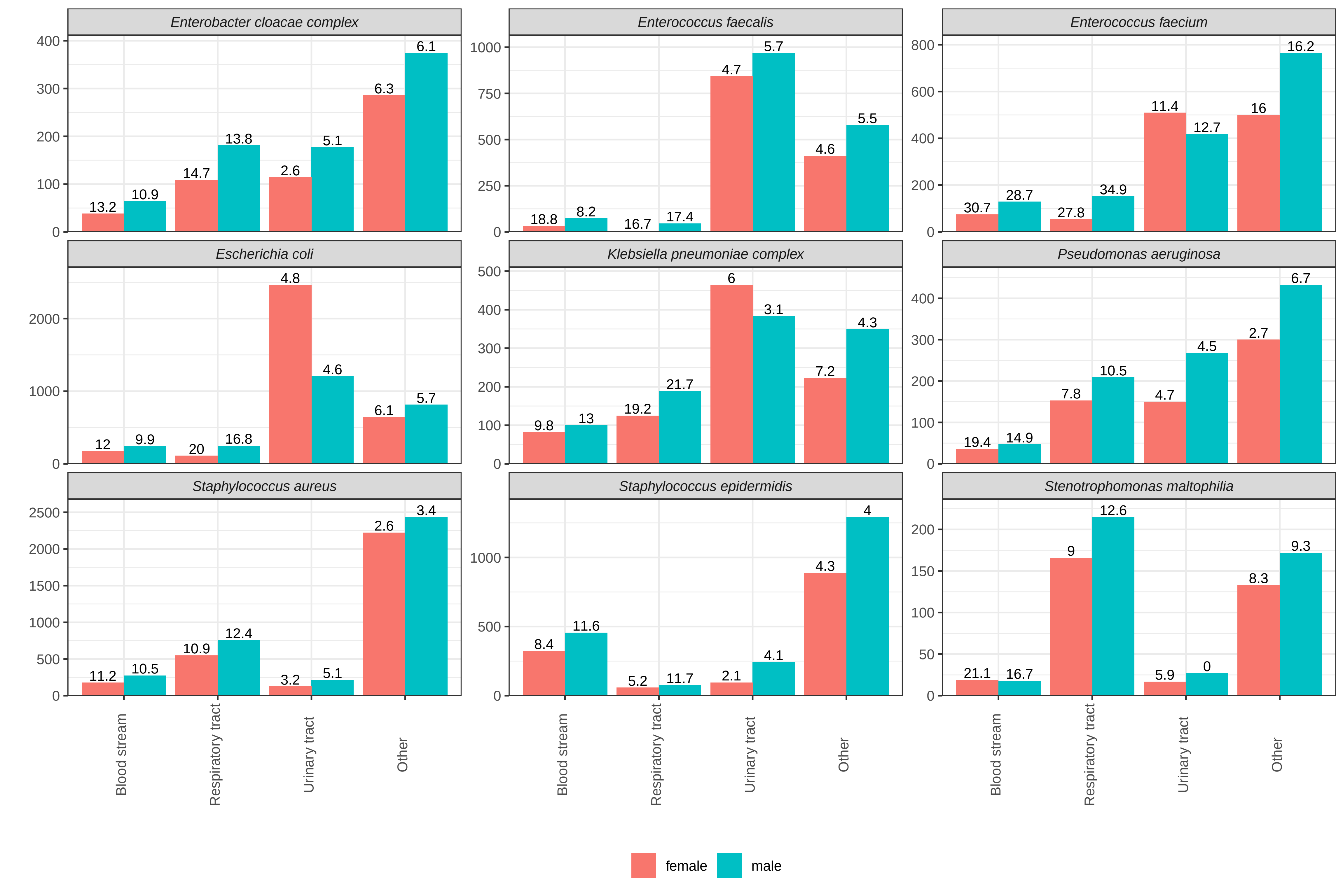

### Supplementary figure 2

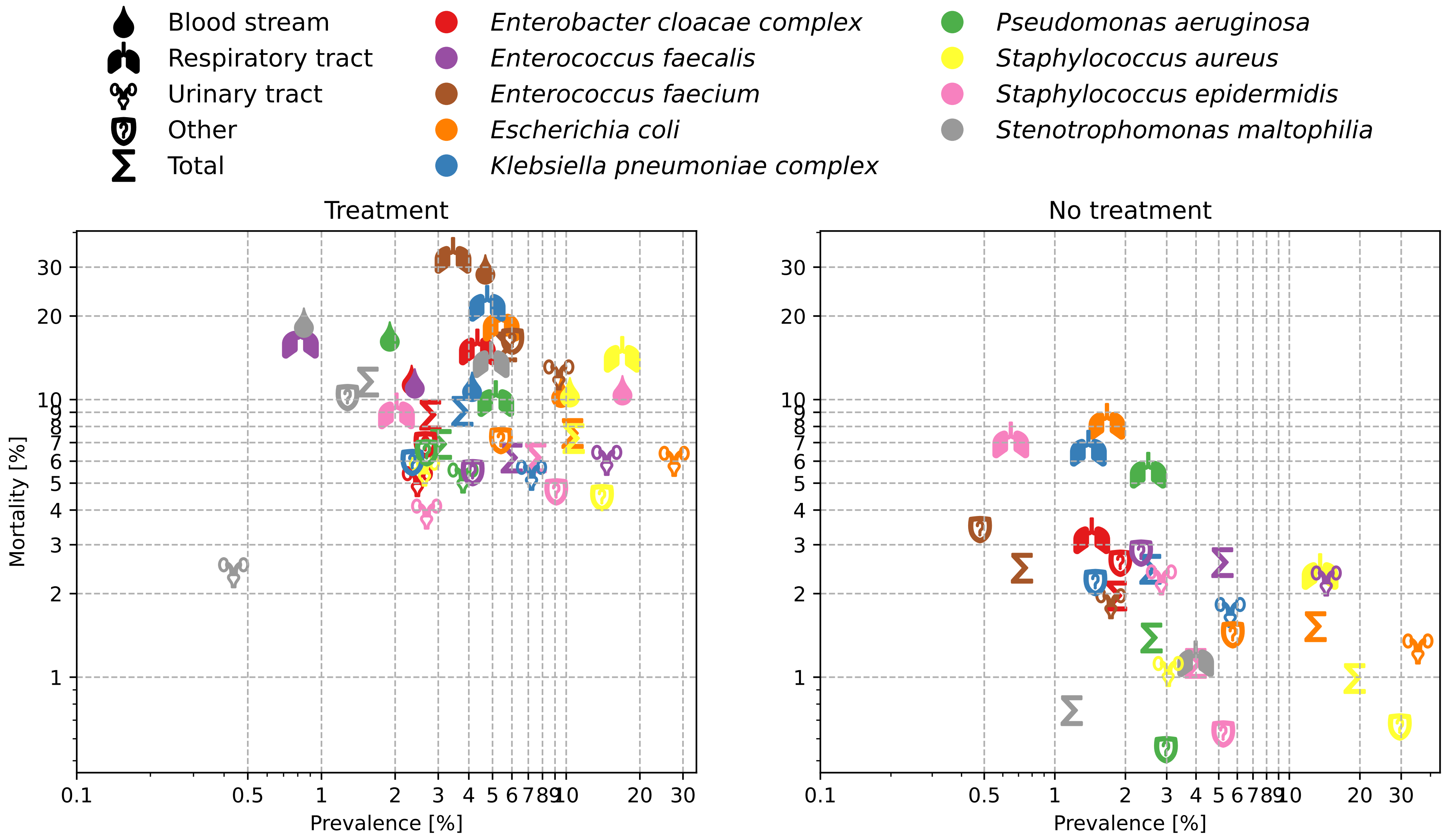

### Supplementary figure 3

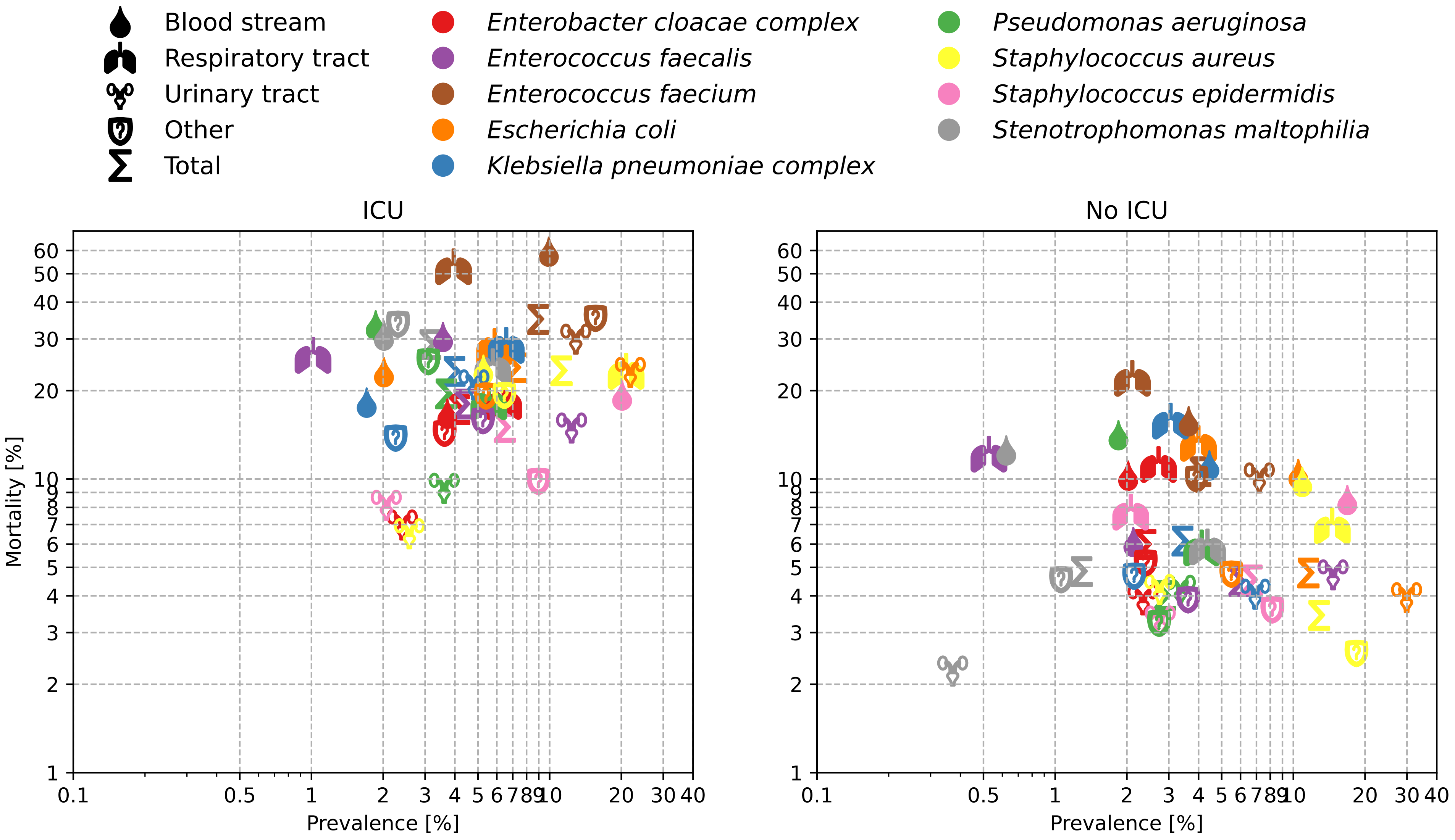

### Supplementary figure 4

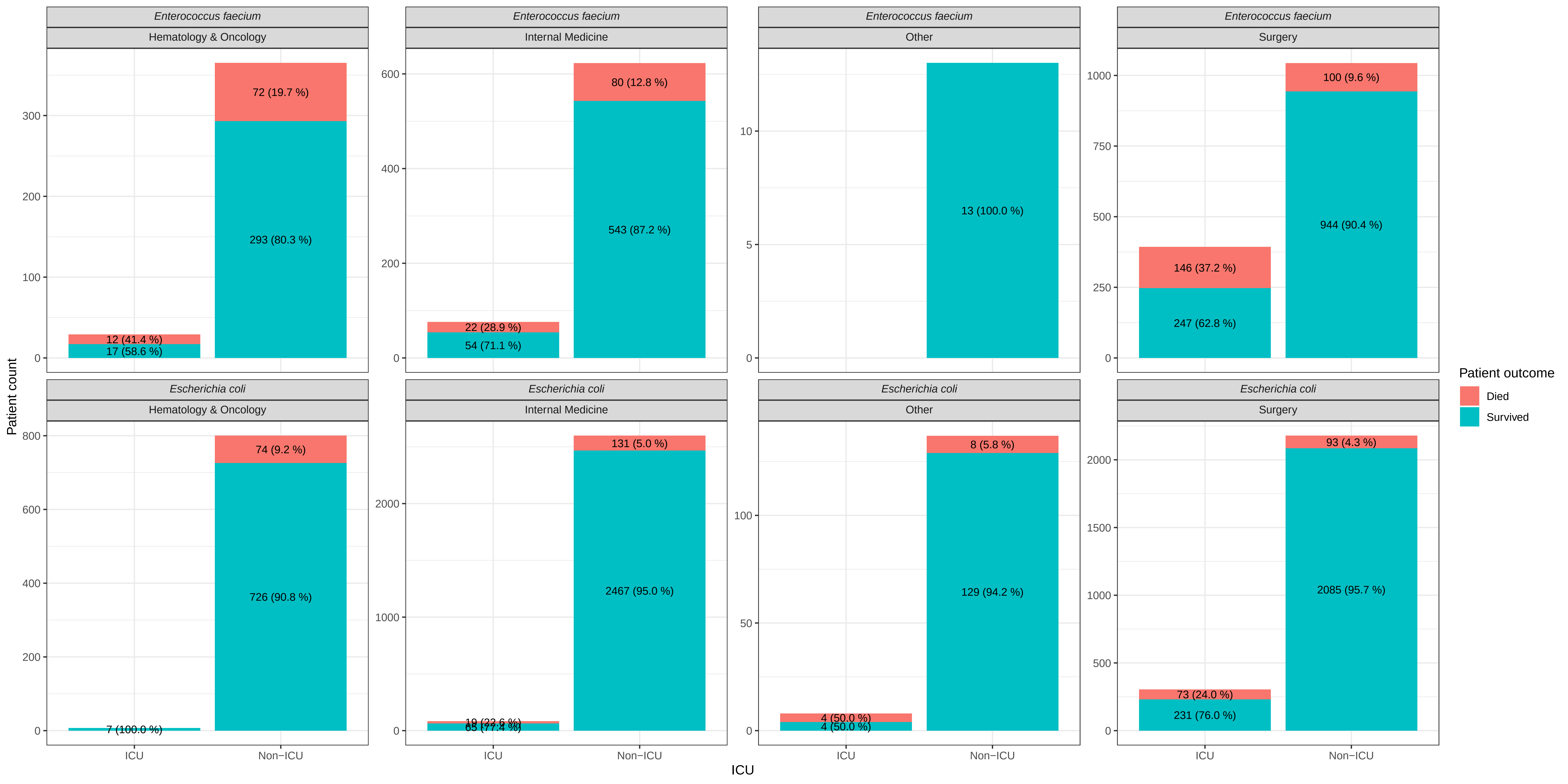
