## Supplementary figure 5 for "Hospitalized patients lack pathogen-specific comorbidity signatures: general vulnerability and case fatality drive differences in mortality"

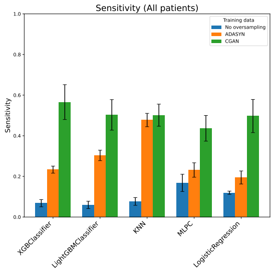


(a)

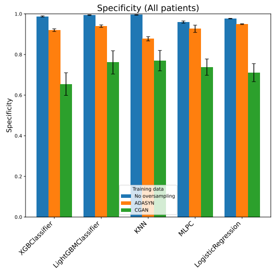


(b)

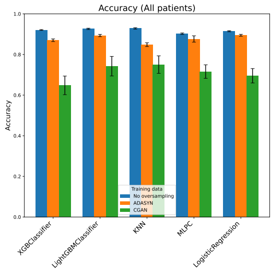


(c)

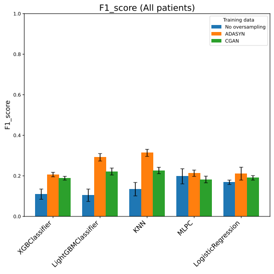


(d)

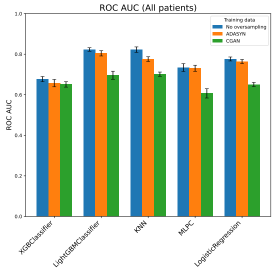


(e)

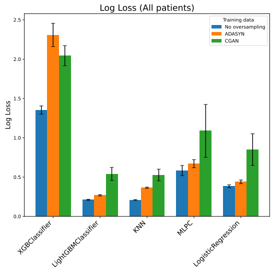


(f)

Supplementary Fig 5: Overall model performance

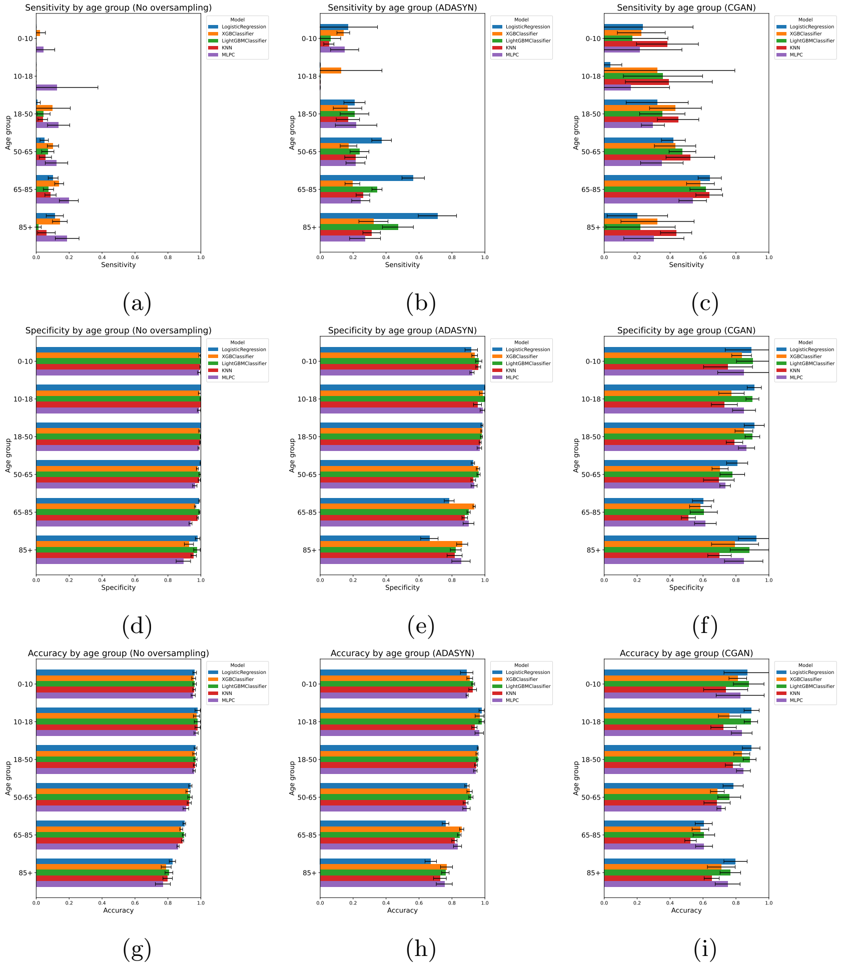


Supplementary Fig 6: Model performance summarized by age groups (Sensitivity + Specificity + Accuracy)

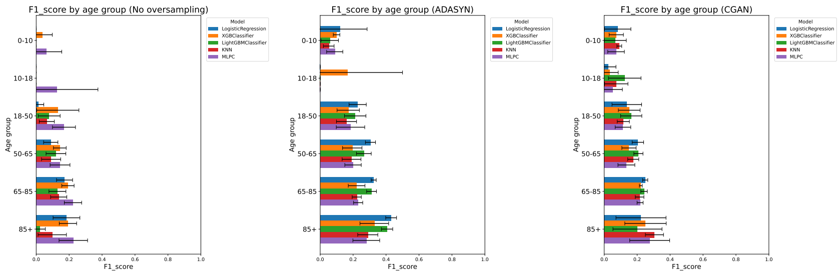


(a) (b) (c)

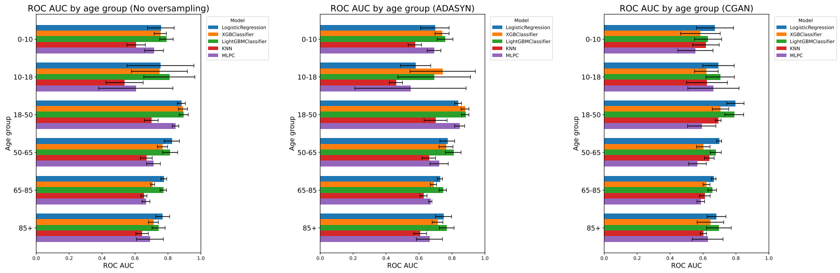


(d) (e) (f)

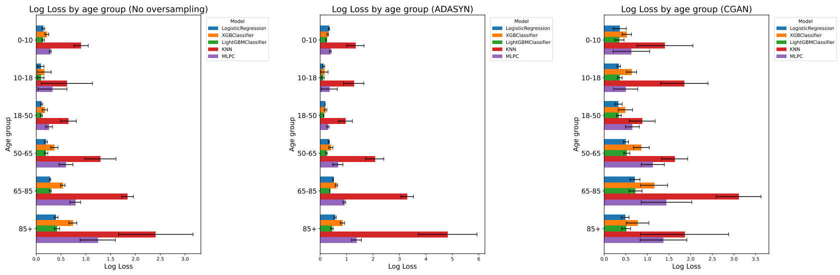


(g) (h) (i)

Supplementary Fig 7: Model performance summarized by age groups (F1-score + ROC AUC + Log Loss)

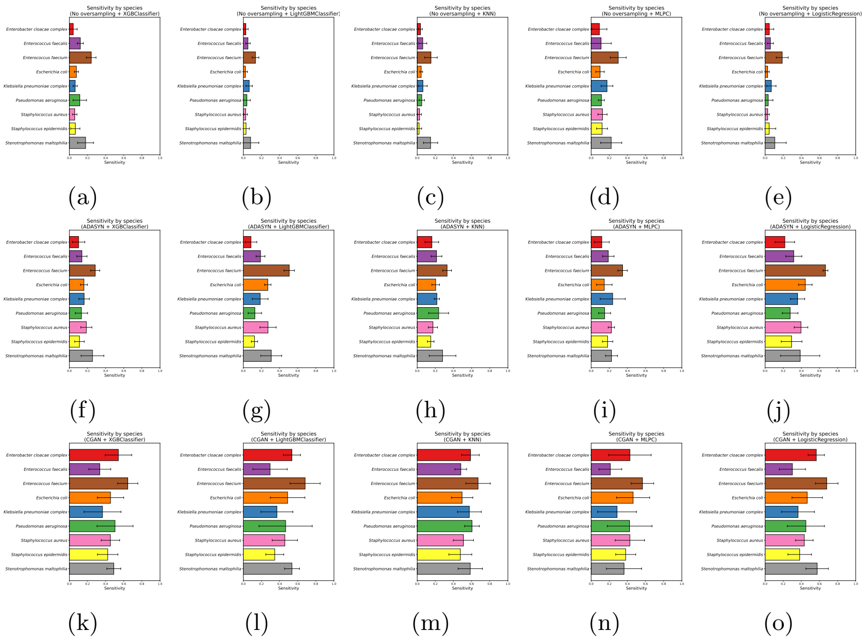


Supplementary Fig 8: Model performance summarized by species: Sensitivity

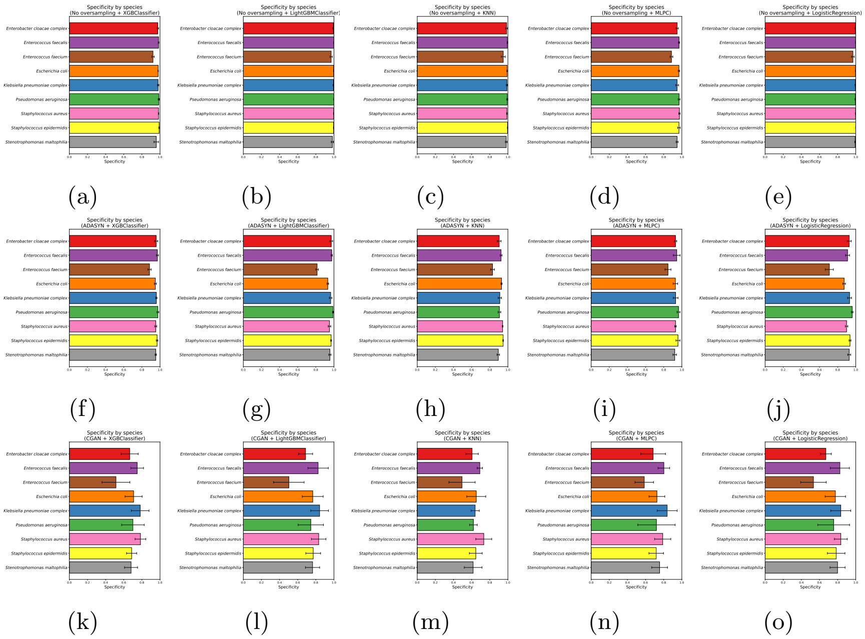


Supplementary Fig 9: Model performance summarized by species: Specificity

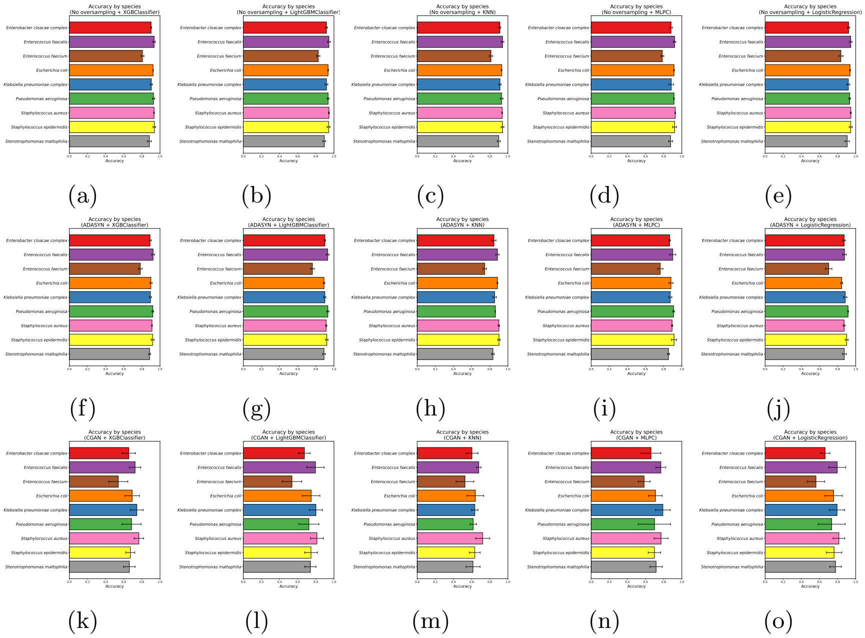


Supplementary Fig 10: Model performance summarized by species: Accuracy

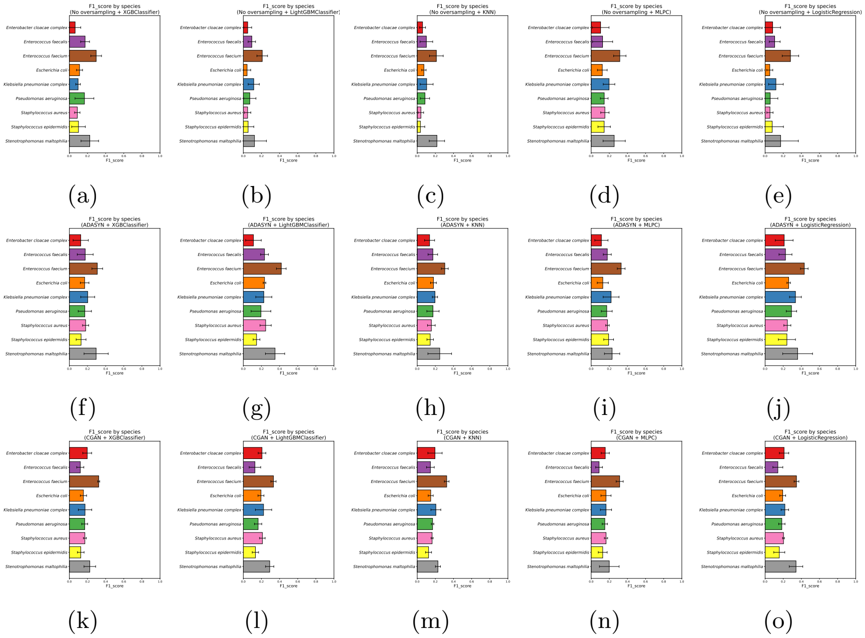


Supplementary Fig 11: Model performance summarized by species: F1-score

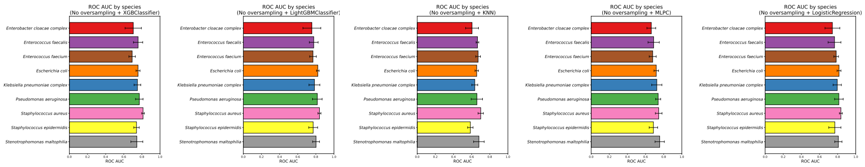


(a) (b) (c) (d) (e)

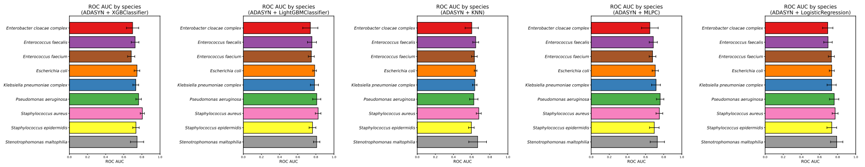


(f) (g) (h) (i) (j)

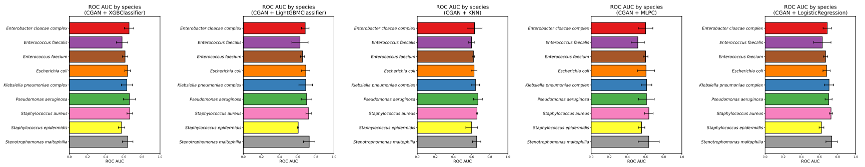


(k) (l) (m) (n) (o)

Supplementary Fig 12: Model performance summarized by species: ROC AUC

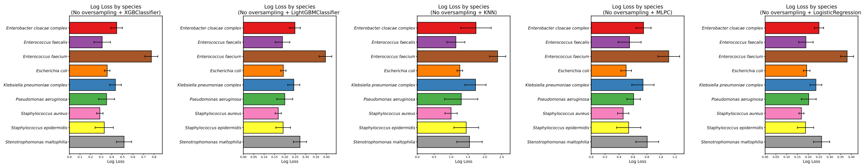


(a) (b) (c) (d) (e)

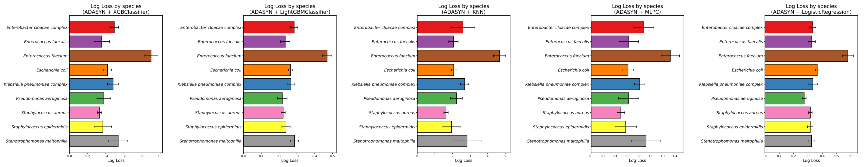


(f) (g) (h) (i) (j)

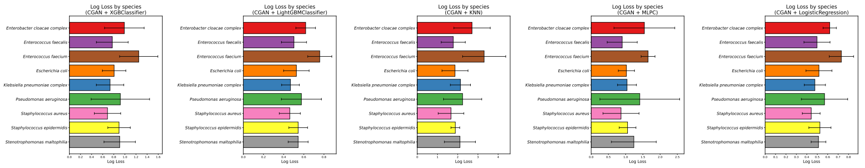


(k) (l) (m) (n) (o)

Supplementary Fig 13: Model performance summarized by species: Log Loss
